# Numerical Cincinnati Stroke Scale versus Stroke Severity Screening Tools for the Prehospital Determination of LVO

**DOI:** 10.1101/2024.05.02.24306794

**Authors:** Holden M. Wagstaff, Remle P. Crowe, Scott T. Youngquist, H. Hill Stoecklein, Ali Treichel, Yao He, Jennifer J. Majersik

**Affiliations:** University of Utah, Department of Emergency Medicine; ESO: Emergency Medical Services Software; Emergency Medicine of Jackson Hole, St. John’s Health; University of Utah, Department of Neurology

**Keywords:** prehospital, emergency medical services, stroke

## Abstract

**Background:** Previous research demonstrated that the numerical Cincinnati Prehospital Stroke Scale (CPSS) identifies large vessel occlusion (LVO) at similar rates compared to a limited number of stroke severity screening tools. We aimed to compare numerical CPSS to additional stroke scales using a national EMS database.

**Methods:** Using the ESO Data Collaborative, the largest EMS database with hospital linked data, we retrospectively analyzed prehospital patient records for the year 2022. Stroke and LVO diagnoses were determined by ICD-10 codes from linked hospital discharge and emergency department records. Prehospital CPSS was compared to the Cincinnati Stroke Triage Assessment Tool (C-STAT), the Field Assessment Stroke Triage for Emergency Destination (FAST-ED), and the Balance Eyes Face Arm Speech Time (BE-FAST). The optimal prediction cut-points for LVO screening were determined by intersecting the sensitivity and specificity curves for each scale. To compare the discriminative abilities of each scale among those diagnosed with LVO, we used the area under the receiver operating curve (AUROC).

**Results:** We identified 17,442 prehospital records from 754 EMS agencies with ≥ 1 documented stroke scale of interest: 30.3% (n=5,278) had a hospital diagnosis of stroke, of which 71.6% (n=3,781) were ischemic; of those, 21.6% (n=817) were diagnosed with LVO. CPSS score ≥ 2 was found to be predictive of LVO with 76.9% sensitivity, 68.0% specificity, and AUROC 0.787 (95% CI 0.722-0.801). All other tools had similar predictive abilities, with sensitivity / specificity / AUROC of: C-STAT 62.5% / 76.5% / 0.727 (0.555-0.899); FAST-ED 61.4% / 76.1%/ 0.780 (0.725-0.836); BE-FAST 70.4% / 67.1% / 0.739 (0.697-0.788).

**Conclusion:** The less complex CPSS exhibited comparable performance to three frequently employed LVO detection tools. EMS agency leadership, medical directors, stroke system directors, and other stroke leaders may consider the complexity of stroke severity instruments and challenges with ensuring accurate recall and consistent application when selecting which instrument to implement. Use of the simpler CPSS may enhance compliance with the utilization of LVO screening instruments while maintaining the accuracy of prehospital LVO determination.

## Background

Early prehospital identification of stroke, coupled with transport to a hospital with appropriate therapeutic capabilities, is a pillar of optimal stroke care (1). For patients experiencing a severe form of stroke known as large vessel occlusion (LVO), endovascular treatment has been shown to reduce morbidity and mortality; yet this treatment is available only at specialized centers (2-5). Frequently, Emergency Medical Services (EMS) clinicians are the first to encounter patients experiencing a stroke (6). It is therefore crucial that EMS clinicians can quickly recognize stroke and possible LVO in a timely manner and transport the patient to a thrombectomy-capable center, when appropriate (7,8).

The optimal screening tool for the identification of LVO by EMS clinicians is an active area of research. The Cincinnati Prehospital Stroke Scale (CPSS), which predates the era of thrombectomy, has been used by many EMS systems to screen patients for stroke. Its high inter-rater reliability and ease of use have led to its widespread implementation in EMS systems around the world (9). For the diagnosis of stroke, the CPSS has a pooled sensitivity of 82.8% and specificity of 56.3% (10). CPSS as a screening tool for identification of LVO was first proposed in 2018 (11). In comparative analyses, the CPSS demonstrated performance comparable to several other scales, such as the Rapid Arterial occlusion Evaluation (RACE), the Los Angeles Motor Scale (LAMS), and the Vision Aphasia Neglect (VAN) scales (12). However, stroke severity tools including the Cincinnati Stroke Triage Assessment Tool (C-STAT) (13), the Field Assessment Stroke Triage for Emergency Destination (FAST-ED) (14), and the Balance Eyes Face Arm Speech Time (BE-FAST) (15), were not included in these evaluations. CPSS and BE-FAST were developed to broadly screen for stroke, while FAST-ED and C-STAT were developed specifically to identify LVO. C-STAT is unique among stroke severity scales in that it was designed to be used after a positive CPSS identified a potential stroke to then identify LVO: similar derivation methods were used to produce the C-STAT as were used for the CPSS (13, 15).

The objective of this study was to compare the predictive characteristics of the numerical CPSS with those of additional stroke severity screening tools: C-STAT, BE-FAST, and FAST-ED.

## Methods

### Study Design and Data Source

We conducted a retrospective observational analysis utilizing prospectively collected, prehospital electronic patient care records (EPCRs) coupled with linked hospital records for encounters from January 1, 2022, to December 31, 2022. These records were sourced from the ESO Data Collaborative research database, a repository of prehospital electronic health record software within the United States. Over 2,000 EMS agencies employing ESO software have opted to participate in the ESO Data Collaborative program, wherein their de-identified records contribute to annual datasets designated for research purposes. Aligned with the National EMS Information System (NEMSIS) version 3.4 standard, the electronic health record system facilitates the comprehensive collection of dispatch, demographic, and clinical attributes pertinent to EMS encounters. Through a distinct health data exchange software, bi-directional data sharing is enabled, allowing for the seamless linkage of emergency department (ED) and hospital data, including International Classification of Diseases (ICD-10) diagnoses, with the EPCR. Participation in the ESO Data Collaborative is voluntary for agencies. All records are de-identified and incorporated into yearly research datasets. No manual data abstraction is conducted; instead, pertinent information is directly extracted from the electronic health record database. Access to these annual research datasets is provided at no charge after undergoing a proposal review process, obtaining institutional review board (IRB) approval, and executing a data use agreement. The IRB at the University of Utah approved of this study, IRB_00161488, and deemed it exempt from human subjects review. This study conforms to Strengthening the Reporting of Observational Studies in Epidemiology (STROBE) guidelines (16).

### Inclusion of Records for Analysis

We included all emergency (9-1-1) EMS calls that involved the use of one or more stroke scales of interest (CPSS, C-STAT, BE-FAST, or FAST-ED) and resulted in transportation to a hospital. We excluded encounters that did not originate from a 9-1-1 call (such as interfacility transports) and those that did not lead to EMS transport to a hospital. We also excluded records that were not linked to ED or hospital diagnosis codes.

### Classification of Stroke and LVO

We used ED and hospital ICD-10 diagnosis codes to determine diagnoses of stroke or transient ischemic attack (TIA) (Supplementary Table 1). We combined hospital and emergency department (ED) diagnoses with ED diagnoses taking precedence; only primary and secondary ICD-10 codes were used. Codes starting with I60, I61, or I62 were classified as intracerebral hemorrhage (ICH). Diagnoses starting with I63 were classified as acute ischemic stroke (AIS). Codes starting with G45 were used to identify transient ischemic attack (TIA). We classified patients who had ICD-10 codes indicating multiple categories of stroke into a separate category of multiple stroke types. While terminological variations exist, large vessel occlusion (LVO) stroke is commonly defined as blockages of the middle cerebral arteries (MCA), internal carotid artery (ICA), and/or basilar artery (BA). We used ICD-10 codes indicating thrombosis or embolism of one or more of these vessels to identify LVO stroke (Supplementary Table 2).

### Stroke Scales

In this study, we analyzed four stroke screening tools: CPSS, C-STAT, BE-FAST, and FAST-ED. Originally, CPSS was unscored, but has since been adapted to a 0-3 scale, assigning one point for each component (face, arm, speech) (11,12). BE-FAST, developed to improve CPSS’s sensitivity and specificity, adds balance and eye components, scoring issues from 0-1 across five criteria (balance, blurry vision, facial droop, arm weakness, speech difficulty), with a total score range of 0-5 (15). However, BE-FAST lacks a designated LVO indication threshold. C-STAT evaluates conjugate gaze (0-2), consciousness level (0-1), and arm weakness (0-1), with scores of 2 or higher suggesting LVO and recommending transport to an endovascular center (13). FAST-ED assesses facial palsy, arm weakness, speech changes, eye deviation, and denial/neglect on a scale from 0-9, with a score of 4 or higher indicating LVO and directing patients to specialized centers (14).

The inclusion of C-STAT as an LVO screening instrument in this study is justified by its unique derivation, by the same research group and methodology as CPSS (13,17). Additionally, it was not included in prior studies regarding numerical CPSS (11,18). C-STAT was conceived as a two-tiered screening strategy, incorporating eye deviation, a clinical marker that has garnered recognition for its predictive value regarding LVOs (18). Similarly, FAST-ED not only assesses eye deviation but also examines patient neglect, a cortical symptom strongly correlated with LVO (14,19).

### Statistical Analysis

We assessed the predictive performance of the CPSS, C-STAT, FAST-ED, and BE-FAST stroke screens by computing the sensitivity and specificity for detecting LVO, along with 95% confidence intervals. Statistically optimal cut-points for LVO detection on each scale were determined by the point where the sensitivity and specificity curves overlapped. The overall discrimination of each LVO scale was assessed using area under the receiver operating characteristic curves (AUROC). A C-statistic (area under the ROC curve) between 0.7 and 0.8 is generally considered acceptable discrimination, while a C-statistic greater than 0.8 is considered excellent discrimination. To determine the presence of statistically significant differences in the discriminative ability of the stroke severity assessment tools, the DeLong method was employed to compare AUROC of CPSS to the other tools. Additionally, within subpopulations where two stroke scales were concurrently applied to the same patient, McNemar’s Chi-squared test with continuity correction was utilized. This approach facilitated a comparative analysis of the sensitivities and specificities. All analyses were conducted using R software (version 4.3.0 (2023-04-21). Power calculation for sample size omitted given the nature of the analysis. Due to the compulsory completion of EMS charts, the issue of missing data is mitigated when conducting statistical analyses.

## Results

### Epidemiology

During the study period, there were a total of 359,064 emergency encounters by 754 EMS agencies with linked hospital data. Of these, 17,442 encounters involving 437 EMS agencies included documentation of one or more of the stroke scales of interest. Among these patients, 5,278 (30.3%) had a linked ICD-10 hospital code indicating a stroke diagnosis. Of the patients diagnosed with stroke, 3,781 (71.6%) were identified with an acute ischemic stroke. Within this group, 837 (21.6%) were diagnosed with a Large Vessel Occlusion (LVO) (Figure 1).

**Figure 1:**
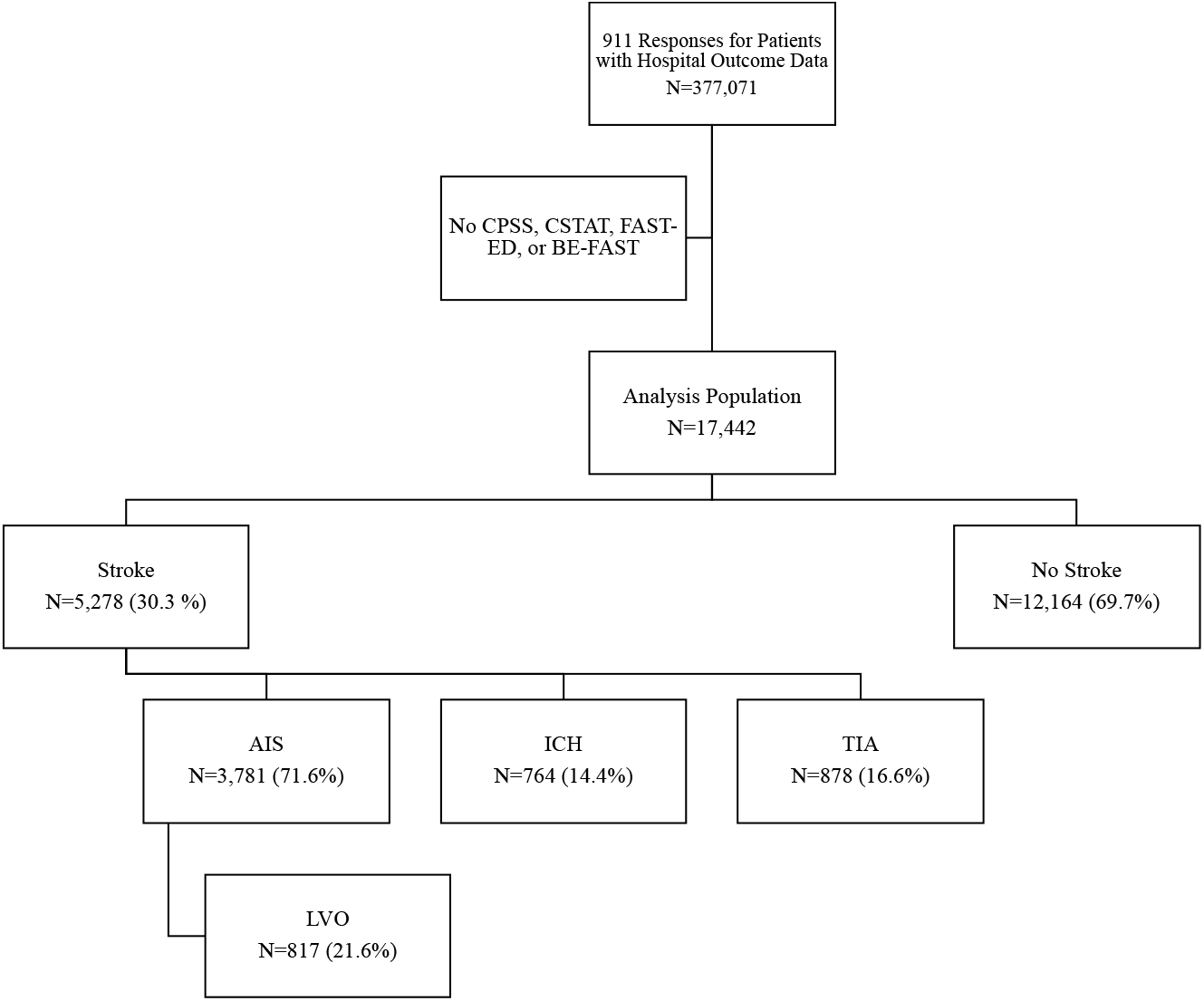
Inclusion of patients for analysis and hospital/emergency department stroke diagnoses. CPSS: Cincinnati Prehospital Stroke Scale; C-STAT: Cincinnati Stroke Triage Assessment Tool; FAST-ED; Field Assessment Stroke Triage for Emergency Destination; BE-FAST: Balance Eyes Face Arm Speech Time; AIS: Acute Ischemic Stroke; ICH: Intracerebral Hemorrhage; TIA: Transient Ischemic Attack

### Patient and Encounter Characteristics

The median age of the patients was 74 years (Interquartile Range: 64-83 years), and 53.5% were female. Most patients (93.6%) were assessed by EMS in urban areas, with 5.3% in rural areas and 1.1% in areas designated as super-rural. The highest certification level of EMS personnel recorded for these cases was Emergency Medicine Technician (EMT) in 5.4% of cases and Paramedic in 92.8%. The Cincinnati Prehospital Stroke Scale (CPSS) was the most frequently used instrument, utilized in 82.5% (n=14,384) of patients, followed by BE-FAST at 10.6% (n=1,845), FAST-ED at 9.8% (n=1,709), and C-STAT at 0.5% (n=89) (Table 1). Details on race and ethnicity are also provided below.

**Table 1.** Patient characteristics stratified by prehospital stroke scale(s) documented. Patient characteristics, including total sample and stratified by stroke scale. Multiple types of stroke scales could be documented per each encounter.

|  | Total<br>N=17442 | CPSS* 82.5%<br>(N=14384) | CSTAT* 0.5%<br>(N=89) | FAST-ED* 9.8%<br>(N=1709) | BE-FAST* 10.6%<br>(N=1845) |
| --- | --- | --- | --- | --- | --- |
| Age, years Median (IQR) | 74 (64-83) | 74 (63-83) | 69 (56-77) | 74 (65-83) | 74 (64-83) |
| Sex, F | 53.6%<br>(9344) | 53.9% (7752) | 56.2% (50) | 53.3% (911) | 52.0% (959) |
| Sex, M | 46.3%<br>(8083) | 46.0% (6617) | 43.8% (39) | 46.7% (798) | 48.0% (886) |
| Sex, Other | 0.0% (15) | 0.0% (15) | 0.0% (0) | 0.0% (0) | 0.0% (0) |
| Stroke Diagnosis, Y | 30.3%<br>(5278) | 30.0% (4313) | 29.2% (26) | 31.2% (533) | 34.3% (633) |
| Hemorrhagic Diagnosis, Y | 4.4%<br>(764) | 4.4% (635) | 9.0% (8) | 5.0% (86) | 3.9% (72) |
| AIS Diagnosis, Y | 21.7%<br>(3781) | 21.3% (3060) | 15.7% (14) | 23.5% (402) | 26.2% (484) |
| TIA Diagnosis, Y | 5.0%<br>(878) | 5.2% (752) | 4.5% (4) | 4.3% (73) | 4.7% (86) |
| LVO Diagnosis, Y | 4.7%<br>(817) | 5.0% (722) | 9.0% (8) | 3.3% (57) | 4.4% (81) |
| Stroke or Tia Impression by EMS, Y | 58.3%<br>(10164) | 57.7% (8306) | 70.8% (63) | 60.0% (1026) | 68.2% (1259) |
| Urban | 93.6%<br>(16328) | 93.2% (13412) | 98.9% (88) | 96.0% (1641) | 93.1% (1718) |
| Rural | 5.3%<br>(916) | 5.4% (777) | 1.1% (1) | 4.0% (68) | 6.3% (116) |
| Super Rural | 1.1%<br>(193) | 1.3% (190) | 0 | 0 | 0.6% (11) |
| EMT, highest pre-hospital provider | 5.4%<br>(935) | 6.3% (911) | 23.6% (21) | 0.5% (8) | 1.1% (21) |
| ALS, highest pre-hospital provider | 92.8%<br>(16187) | 91.5% (13165) | 75.3% (67) | 98.3% (1680) | 98.8% (1823) |
| Race/Ethnicity: White, non-Hispanic | 67.2%<br>(11715) | 68.8% (9901) | 64.0% (57) | 62.7% (1072) | 62.1% (1146) |
| Race/Ethnicity: Black, non-Hispanic | 15.3%<br>(2669) | 16.2% (2326) | 29.2% (26) | 17.6% (300) | 4.2% (77) |
| Race/Ethnicity: Asian, non-Hispanic | 1.4%<br>(247) | 1.4% (203) | 2.2% (2) | 1.2% (21) | 1.4% (26) |
| Race/Ethnicity: Hispanic | 5.5%<br>(956) | 5.2% (752) | 1.1% (1) | 3.1% (53) | 10.0% (185) |
| Race/Ethnicity: Other, non-Hispanic | 10.6%<br>(1855) | 8.3% (1202) | 4.5% (4) | 15.3% (263) | 22.3% (411) |

### Predictive Performance of Stroke Screening Instruments

Table 2 breaks down the performance of each stroke scale across different cut points. Figure 2 displays the AUROC curves for each of the stroke severity tools. The performance of CPSS was maximized at a cut point of 2 or higher, achieving a sensitivity of 76.9%, a specificity of 68.0%, and an AUROC of 0.787 (95% CI 0.722-0.801). C-STAT demonstrated maximum performance at a cut point of ≥2, with a sensitivity of 62.5%, a specificity of 76.5%, and an AUROC of 0.727 (95% CI 0.555-0.899). BE-FAST showed its best performance at a cut point of ≥3, with a sensitivity of 70.4%, a specificity of 67.1%, and an AUROC of 0.739 (95% CI 0.697-0.788). FAST-ED achieved its optimal performance at a cut point of ≥4, with a sensitivity of 61.4%, a specificity of 76.1%, and an AUROC of 0.780 (95% CI 0.725-0.836). Although CPSS had the highest AUROC, it did not show statistically significant discriminative abilities compared to C-STAT (p=0.689), BE-FAST (p=0.727), and FAST-ED (p=0.269), according to DeLong’s test for two correlated receiver operating curves. When comparing different subpopulations where CPSS and an additional stroke severity assessment tool were used, we analyzed sensitivity and specificity using McNemar’s Chi-square test. The numerical CPSS outperformed C-STAT (p<0.001) and FAST-ED (p<0.001); however, there was no statistical difference between numerical CPSS and BE-FAST (p=0.212).

**Table 2.** Predictive performance of the CPSS, CSTAT, FAST-ED, and BE-FAST screening instruments for detecting LVO stroke.

\*Optimal cut-point identified using sensitivity and specificity curves from the present study.
|  | Sensitivity | Specificity | PLR | NLR | Area under ROC curve,<br>C-Statistic (95% CI) |
| --- | --- | --- | --- | --- | --- |
| CPSS (N=13993) |  |  |  |  | 0.787 (0.772-0.801) |
| ≥1 | 0.943 | 0.463 | 1.758 | 0.123 |  |
| ≥2* | 0.769 | 0.68 | 2.402 | 0.34 |  |
| 3 | 0.56 | 0.84 | 3.497 | 0.524 |  |
| CSTAT (N=89) |  |  |  |  | 0.727 (0.555-0.899) |
| ≥1 | 0.875 | 0.469 | 1.648 | 0.266 |  |
| ≥2 † | 0.625 | 0.765 | 2.664 | 0.49 |  |
| ≥3 | 0.5 | 0.864 | 3.682 | 0.579 |  |
| 4 | 0 | 0.901 | 0 | 1.11 |  |
| FAST-ED<br>(N=1604) |  |  |  |  | 0.780 (0.725-0.836) |
| ≥1 | 0.965 | 0.335 | 1.452 | 0.105 |  |
| ≥2 | 0.912 | 0.492 | 1.794 | 0.178 |  |
| ≥3 | 0.737 | 0.652 | 2.117 | 0.404 |  |
| ≥4 † | 0.614 | 0.761 | 2.568 | 0.507 |  |
| ≥5 | 0.526 | 0.85 | 3.506 | 0.557 |  |
| ≥6 | 0.386 | 0.909 | 4.251 | 0.675 |  |
| ≥7 | 0.263 | 0.953 | 5.574 | 0.773 |  |
| ≥8 | 0.14 | 0.977 | 6.102 | 0.88 |  |
| 9 | 0.018 | 0.996 | 4.83 | 0.986 |  |
| BE-FAST<br>(N=1768) |  |  |  |  | 0.739 (0.691-0.788) |
| ≥1 | 0.951 | 0.293 | 1.345 | 0.168 |  |
| ≥2 | 0.889 | 0.483 | 1.719 | 0.23 |  |
| ≥3* | 0.704 | 0.671 | 2.14 | 0.441 |  |
| ≥4 | 0.444 | 0.804 | 2.266 | 0.691 |  |
| 5 | 0.222 | 0.942 | 3.843 | 0.826 |  |

**Figure 2:**
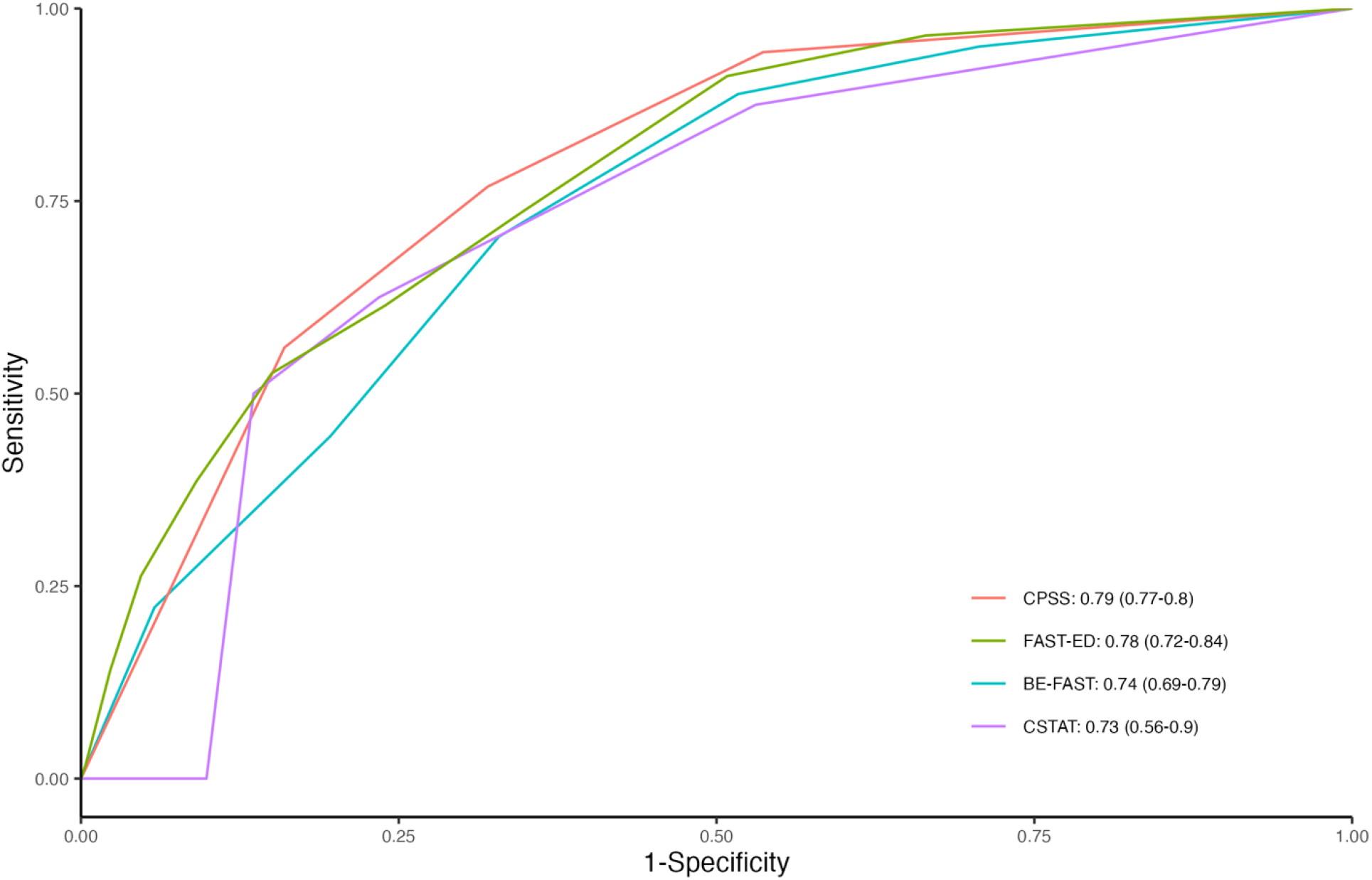
ROC curves for prehospital stroke severity scales. CPSS: Cincinnati Prehospital Stroke Scale; C-STAT: Cincinnati Stroke Triage Assessment Tool; FAST-ED; Field Assessment Stroke Triage for Emergency Destination; BE-FAST: Balance Eyes Face Arm Speech Time

## Discussion

Our retrospective analysis, drawing upon a robust nationwide EMS database comprising over 17,400 patient records with linked hospital outcomes, demonstrated that the simpler, numerical CPSS achieved LVO identification performance comparable to that of the more complex C-STAT, FAST-ED, and BE-FAST stroke severity screening tools. Despite lower AUROC values for C-STAT, FAST-ED, and BE-FAST compared to CPSS, these differences were not statistically significant in terms of their capacity for prehospital LVO detection. However, in subpopulations where CPSS and either C-STAT or FAST-ED were employed, CPSS exhibited superior performance.

C-STAT was initially developed from two extensive tPA trial datasets, targeting severe strokes—defined as those with a National Institutes of Health Stroke Scale (NIHSS) score of 15 or higher—using regression analysis to identify key predictors of severe stroke (13). Subsequent validation assessed the tool’s ability to detect severe and moderate strokes as well as LVOs. This approach was questioned as only a minority of patients fell into the “severe” stroke category. Moreover, the tool’s prospective validation in the prehospital setting involved fewer than 60 patients and conducted among clinicians who had not received formal training (21). The sensitivity, specificity, and AUROC of C-STAT in this validation study were higher than those observed in broader database studies, including the PRESTO trial and our current research (18).

FAST-ED was validated using the STOPStroke dataset, which comprised patients suspected of having an ischemic stroke and who underwent imaging within 24 hours of potential stroke onset (14). The creators of FAST-ED chose five NIHSS components most closely associated with LVO for the scale. Research associates retrospectively utilized the physical exam results and NIHSS scores recorded by admitting neurologists to determine the operating characteristics of FAST-ED on the detection of LVO in this cohort. In practical prehospital applications, however, the interrater reliability among clinicians was recorded at 0.66, indicating challenges with recall and real-world utilization of the test (22). In contrast, CPSS demonstrated higher interrater reliability, scoring 0.83 between EMS clinicians and neurology attendings, which suggests better recall and application by prehospital clinicians (23).

In many EMS systems, a positive result from CPSS or a similar stroke screening tool typically triggers a secondary evaluation for LVO, following the American Heart Association/American Stroke Association (AHA/ASA) Mission Lifeline guidelines (24). This step is critical for deciding the optimal transport destination of patients suspected of having a stroke. Despite its critical importance, stroke, including LVO, is a relatively rare event for individual clinicians. Our study, consistent with previous research, did not identify a superior stroke severity tool for prehospital LVO detection. However, given the similar effectiveness of these tools, EMS agencies and their decision-makers should consider factors like training burden, integrated reliability, and implementation costs when choosing a screening tool. Furthermore, it is essential to consider the ability of providers to accurately recall and appropriately apply stroke severity tools, particularly given the stressful and varied practice environments faced by EMS clinicians. As CPSS is widely used and requires less training, its continued implementation, coupled with ongoing education, could be less burdensome.

Our work aligns with previous findings that the numerical CPSS effectively identifies LVO and compares favorably to other stroke severity tools, with a cut point of 2 showing optimal statistical performance (11, 16). The PRESTO study in the Netherlands, which prospectively evaluated various prehospital stroke scales, also found CPSS as effective as eight other tools (17). Despite no significant statistical differences in operational characteristics among the scales, the RACE scale might slightly outperform numerical CPSS. A significant limitation affecting the external validity and generalizability of this study is the variability in EMS training between the U.S. and the Netherlands. In the Netherlands, paramedics start their careers as experienced nurses and undergo extensive training, whereas in the U.S., the path to becoming a paramedic begins with a basic EMT certification, which requires approximately 120 hours of training, followed by an additional 1200 to 1800 hours to advance to paramedic status.

## Limitations

Our retrospective analysis relied on existing electronic patient care records, introducing potential risks of incomplete or inaccurate data. Variability in data quality across EMS agencies and regions could affect the reliability of our conclusions. Moreover, using ICD-10 codes for stroke and LVO diagnosis introduces potential errors due to variable coding practices among institutions and possible discrepancies influenced by coders’ proficiency and changing coding criteria. However, this is a standard research method that often yields acceptable results when using primary and secondary codes (25). Our study, limited to 2022, may not reflect changes in EMS practices, technological advancements, or clinical guideline updates that could affect stroke screening tool performance. It also did not examine differences in interrater reliability among EMS providers using various tools, nor the impact of EMS personnel’s proficiency on screening effectiveness, highlighting areas for future research. Additionally, while previous research suggests adding visual and balance assessments could improve detection of posterior circulation strokes—a critical and debilitating subset of LVOs—our study did not explore screening tool performance by LVO or stroke locations (26). Further research is needed to assess whether these assessments significantly enhance detection and affect patient routing and management in a prehospital setting.

## Conclusions

The findings of our study, and totality of evidence thus far, suggest that numerical CPSS performs as well as more complex stroke severity tools. When determining which screening instrument to implement, EMS agency leadership, medical directors, stroke system directors, and other healthcare decision-makers should consider the training requirements to maintain proficiency and consistency in application. Consideration of using numerical CPSS, as opposed to more complicated stroke severity tools with associated increased investment of time and resources, should be considered. Our findings contribute to the evolving landscape of stroke care and continued refinement of current protocols and the strategic allocation of EMS resources.

## Data Availability

All data supporting the findings of this study are available within the article and its supplementary materials. Further inquiries can be directed to the corresponding author.

## Acknowledgements

We express our gratitude to all participating EMS agencies and their personnel who contributed data to the ESO Data Collaborative, making this study possible. Thanks to the ESO Data Collaborative team for their assistance in data management and provision, ensuring that we had a robust dataset for our analysis. We extend our gratitude to our colleagues who support and inspire us in innumerable ways.

